# A Prospective Study on the Financial Impact of Breast Cancer Treatment over a One-Year Period for Patients at a Rural Cancer Center

**DOI:** 10.1101/2025.05.07.25326315

**Authors:** Christopher D. Manko, Khadija Khan, Tyler Zlupko, Nathan Witteveen, Emily Thresher, Jennifer Panek, Apurva Inamdar, Cynthia Perry-Keaty

## Abstract

**Purpose:** Patients undergoing breast cancer treatment experience financial burden that can lead to stress and affect access to care. This study investigates the financial burden patients with breast cancer face.

**Methods:** Change in level of self-reported financial burden was evaluated by determining the difference of the Comprehensive Score for Financial Toxicity-Functional Assessment of Chronic Illness Therapy (COST-FACIT). The survey was administered at baseline, 3, 6, and 12 months to patients undergoing treatment within a rural cancer center to understand patterns with financial burden. A higher score indicated less financial toxicity. Health insurance literacy data (at baseline and 1 year) and support services data (at baseline, 3, 6, and 12 months) were also collected through survey distribution. Demographic data were also collected.

**Results:** COST-FACIT scores significantly increased over the course of the year (p < 0.001) and for patients at or above the age of 65 years (p = 0.001). There was no statistical difference between insurance type (private or public) or between the stages of breast cancer.

**Conclusion:** COST-FACIT scores were higher with treatment over time and if the patient was 65 or over.

**Implication for Cancer Survivors:** Additional financial support should be provided to patients, especially early on in their cancer treatment, with special focus given to patients under the age of 65. This support could help effectively mitigate financial burden.

## Introduction

Financial toxicity is a metric that quantifies financial hardship. It is defined as the burden that can occur due to direct and indirect medical costs [1, 2]. A systematic review by Smith et al., found that 49% patients with cancer had financial toxicity [3]. Prior work has demonstrated that the financial toxicity is not evenly distributed. One cross-sectional study found that female survivors of breast or gynecological cancer of lower age, lower income, and less education were found to experience worse financial toxicity [4]. While many services exist to help patients cope with financial issues, there may be barriers to accessing or utilizing these services.

The second most common cause of cancer for women in the United States is breast cancer [5]. In 2020 alone, over 2 million new cases were diagnosed globally [6]. A diagnosis of breast cancer or recurrence can have a significant financial impact on patients and their families. Throughout cancer treatment, the financial burden of breast cancer may accrue through expenses such as deductibles and copayments. Those diagnosed with metastatic breast cancer can incur compounded costs of indefinite treatment [7]. Patients report feelings of fear and stress in anticipating long-term costs associated with breast cancer [7]. Additionally, costs may impact the treatment plan. In one study utilizing an 88-item web-based questionnaire developed with patient advocates, 43% of surveyed women reported that cost was a factor that influenced decisions surrounding treatment options [8]. Due to the established financial toxicity of breast cancer diagnoses, it becomes imperative to supply breast cancer patients with support services and continually assess the effectiveness and utilization of these services in alleviating financial burden.

The Guthrie Cancer Center is a rural cancer center that provides treatment for breast cancer. Breast cancer care at Guthrie Cancer Center includes a supportive multidisciplinary team with assistance available for financial concerns related to breast cancer treatment. This research study aims to better understand the financial impact that a breast cancer diagnosis has on patients at Guthrie, and to assess how well the existing support services are utilized.

## Methods

This study was a prospective, single-arm, longitudinal, cohort study. Subjects diagnosed with breast cancer (new diagnosis or recurrent) within the past 2 years were identified by a review of the Epic electronic medical record and review of provider schedules. Data collected from the review of electronic medical records included type of health insurance, demographic data, and information related to patients’ breast cancer such as date of diagnosis and stage of progression. Inclusion criteria were subjects 18 years and older diagnosed with breast cancer or a recurrence of breast cancer within the past 2 years. Additionally, patients must have been willing to sign consent and able to read and complete surveys in English. Eligible subjects were invited to participate in the study by a research team member at an already scheduled visit to the Guthrie Cancer Center. The study aimed to collect data for 100 subjects.

The patient-reported financial impact of the breast cancer diagnosis was surveyed with the Comprehensive Score for Financial Toxicity-Functional Assessment of Chronic Illness Therapy (COST-FACIT) [9]. This tool has previously demonstrated valid reliable results when measuring financial toxicity associated with cancer patients [10]. A higher COST-FACIT score indicates a larger level of financial well-being for cancer patients.

The usage of a Guthrie support services survey was also utilized. The COST-FACIT (Version-2) and a study-specific Support Services Survey were administered at four time points: baseline (within 60 days of a new diagnosis), 3 months, 6 months, and 1 year. The Health Insurance Literacy Measurement (HILM) was utilized and administered at baseline and 1 year to better understand patient confidence and proactiveness with insurance [11]. A lower score indicates greater confidence when working with health insurance. This was tested for significance with a paired samples t-test.

The COST-FACIT survey was scored using the associated scoring guidelines [9]. COST-FACIT scores were first analyzed against the time of survey completion. ANOVA with Greenhouse-Geisser correction and post hoc pairwise comparisons with Tukey’s adjustment were performed to determine statistically significant differences for scores versus time. The stages of cancer, age, and insurance type were seen as potential confounders. They were tested directly to better understand their relationship with the perceived financial burden. Scores were analyzed against stage of breast cancer using the one-way ANOVA. COST-FACIT scores were then tested using an independent samples t-test against age (at and over 65 versus under 65). Finally, an independent samples t-test was performed to determine any differences between public versus private insurers for patients versus COST-FACIT scores. Statistical significance is defined as p < 0.05. The study is approved by The Guthrie Clinic Institutional Review Board and registered on ClinicalTrials.gov.

## Results

103 patients were enrolled in the study with 1 ineligible at the beginning. 87 (84.5%) completed the 12-month COST-FACIT survey. 51 of the 103 (49.5%) patients had private insurance whereas the other 52 had public insurance which included both Medicare and Medicaid. Table 1 depicts the breakdown of the eligible patient demographic information. Data was collected with the first survey completion on 09/04/2018 to the last one on 06/15/2022.

**Table 1.** Demographics for eligible patient cohort. Number (N) provided for all variables. Age, stage, and insurance types are also provided as percentages. HILM and COST-FACIT scores are also provided as mean and standard deviation.

| Variable | N | Descriptive |
| --- | --- | --- |
| Age at Baseline | 102 |  |
| < 65 | 71 | 69.6% |
| >= 65 | 31 | 30.4% |
| Staging |  |  |
| I | 41 | 40.2% |
| II | 31 | 30.4% |
| III | 18 | 17.6% |
| IV | 12 | 11.8% |
| Insurance |  |  |
| Private | 50 | 49% |
| Government | 52 | 51% |
| HILM |  |  |
| Baseline | 99 | 9.76 (5.79) |
| 1-Year | 73 | 8.19 (4.86) |
| COST-FACIT |  |  |
| Baseline | 102 | 22.1 (9.2) |
| 3-Months | 90 | 23.6 (8.84) |
| 6-Months | 86 | 25.2 (7.76) |
| 1-Year | 87 | 25.2 (7.62) |

Patient HILM scores significantly decreased between baseline and 1 year (p = 0.018). This indicates that health insurance confidence and proactiveness improved for patients over the course of the year. For the support services survey, it appeared that a majority of patients either did not use and/or were not interested in using the services that the cancer center had to offer (figures 1-4). Overall, at baseline, discussing financial concerns with an oncology social worker was the most utilized service (n=29) and continued to be the most utilized at 1 year. However, the number of patients discussing financial concerns decreased (n = 16). 101 participant responses were reported at baseline and 85 in the 1-year survey. Figure 5 illustrates the number of participants at the beginning and end of the study.

**Figure 1.**
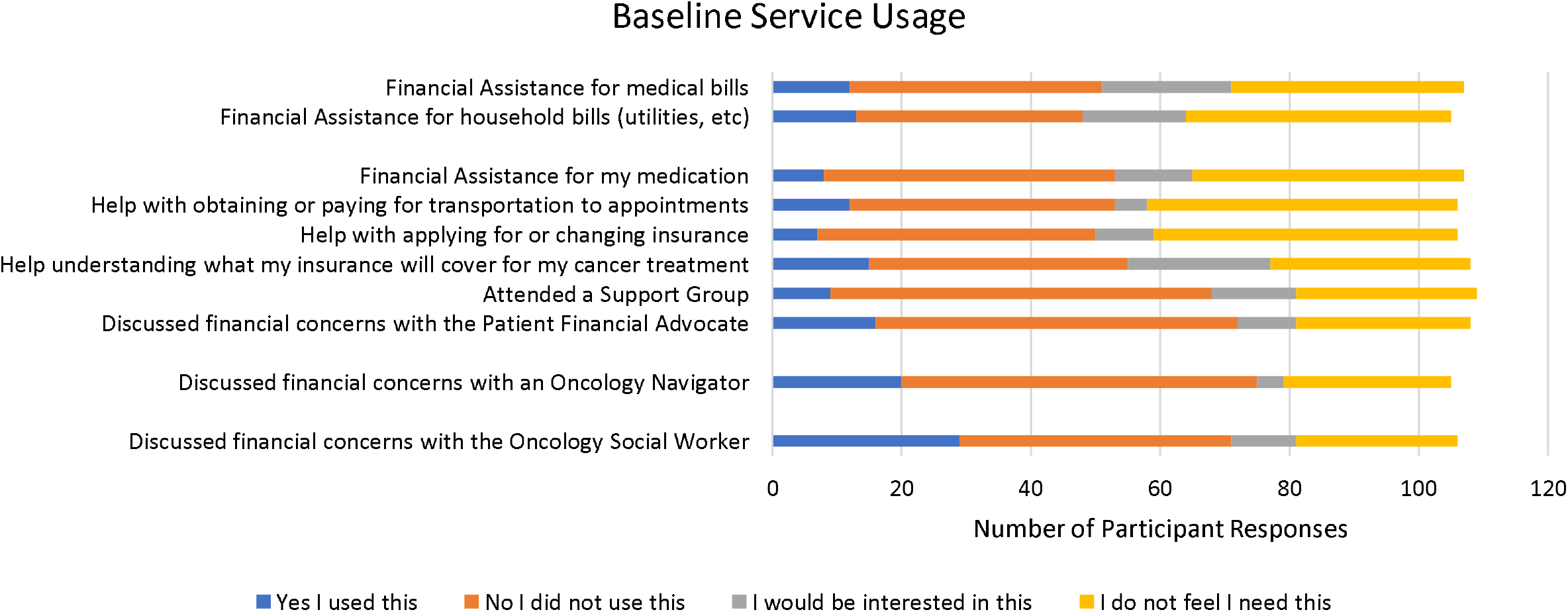
Baseline Support Services Survey.

**Figure 2.**
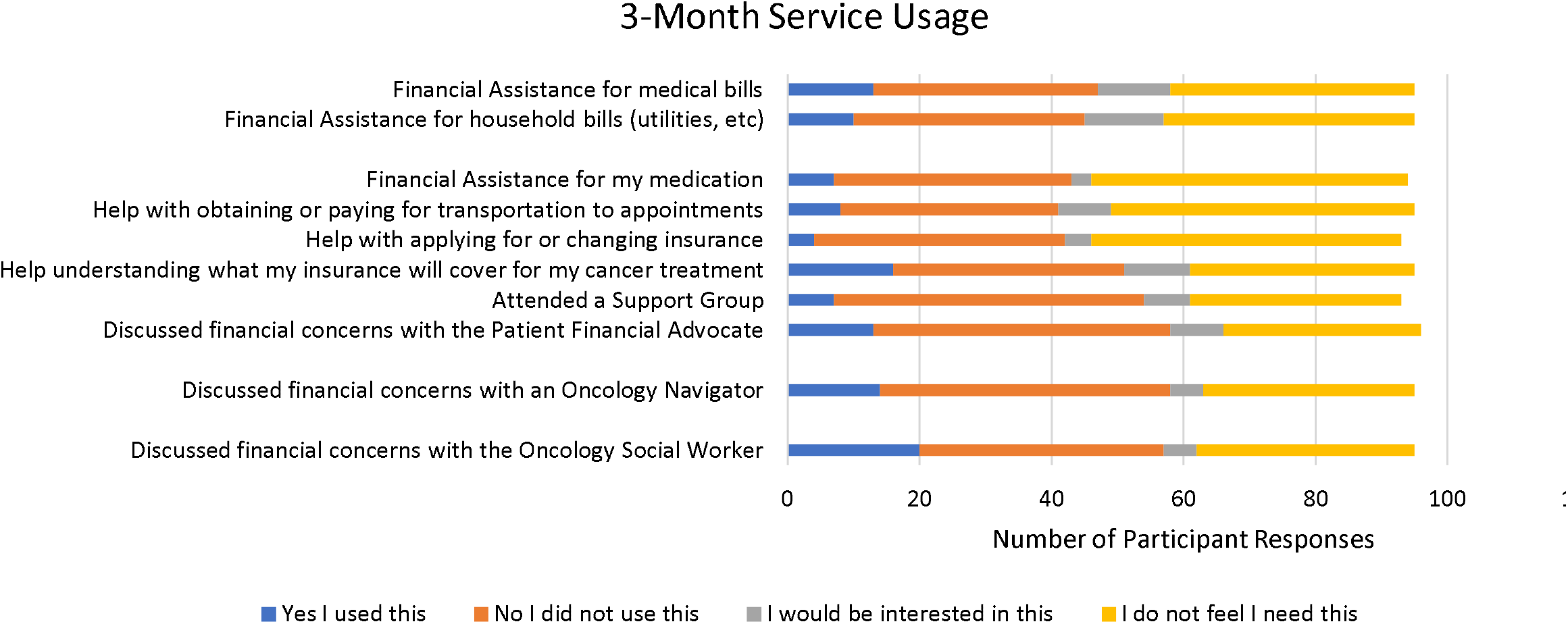
3-Month Support Services Survey.

**Figure 3.**
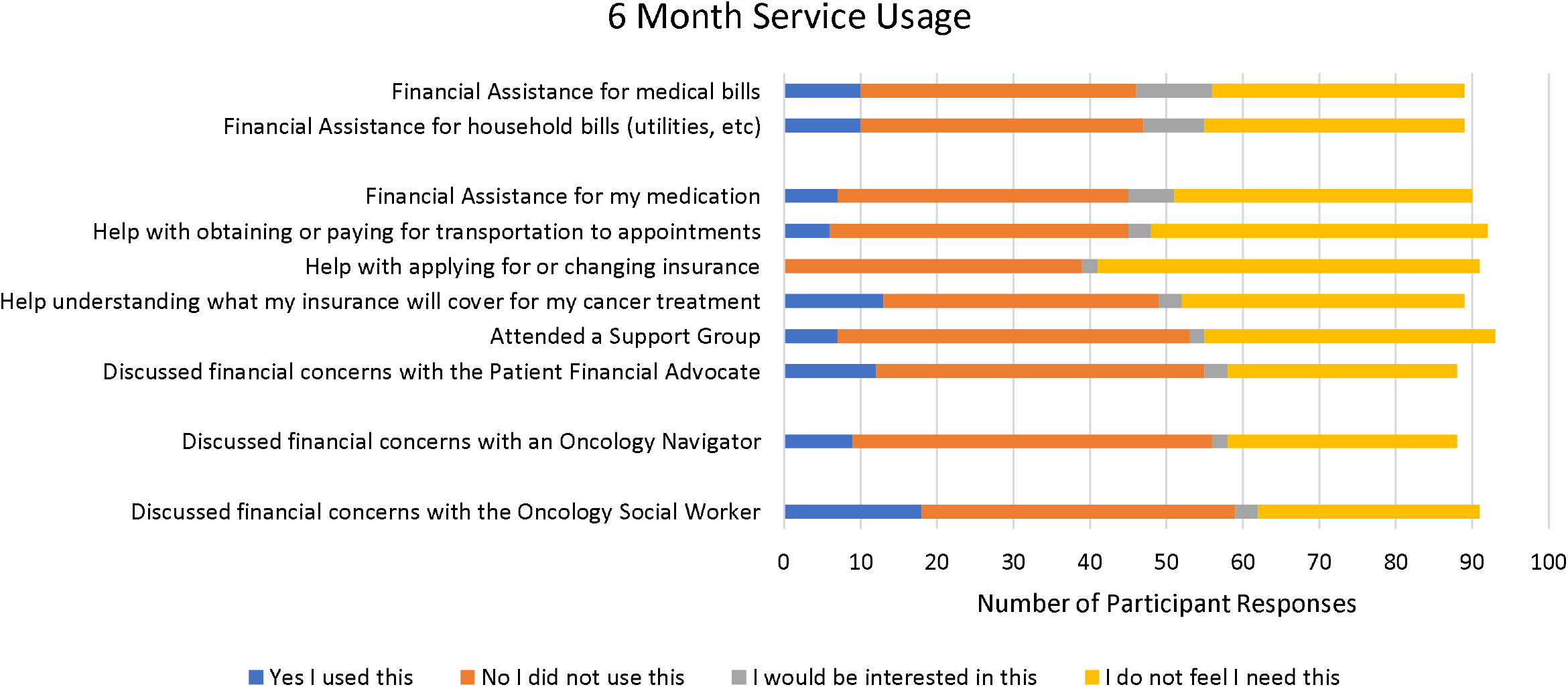
6-Month Support Services Survey.

**Figure 4.**
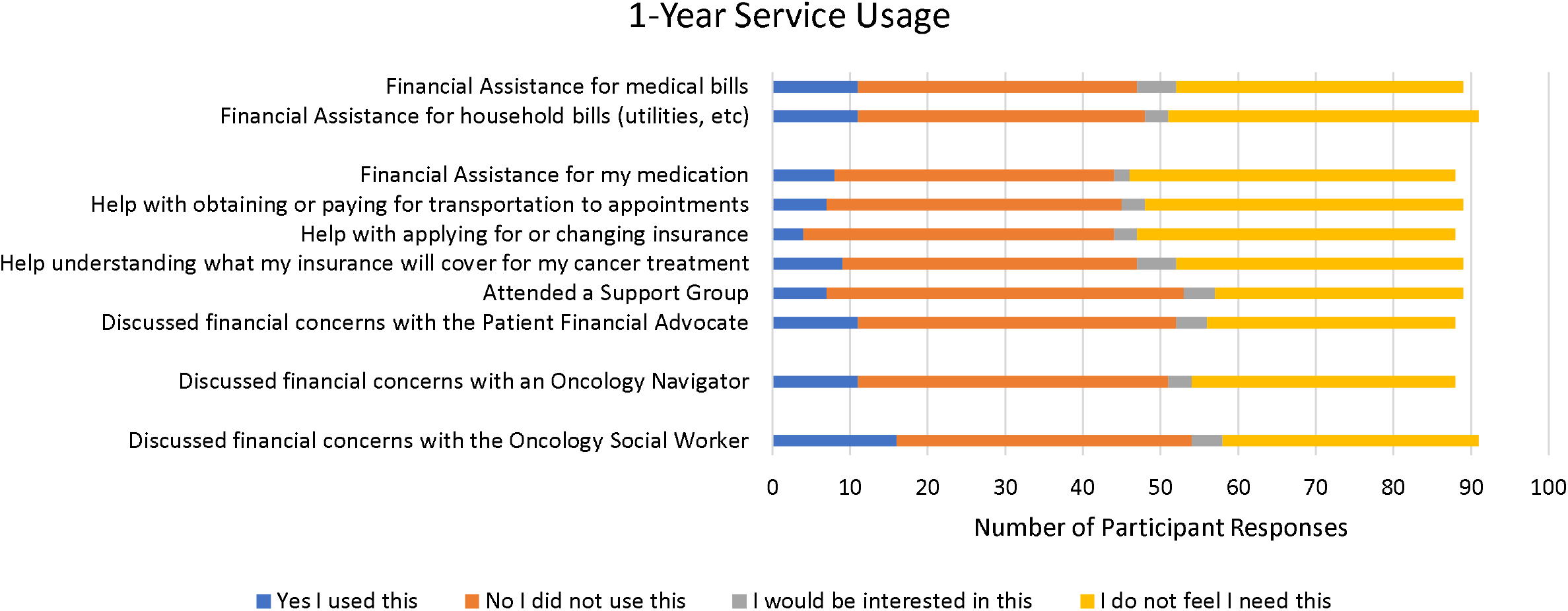
1-Year Support Services Survey.

**Figure 5.**
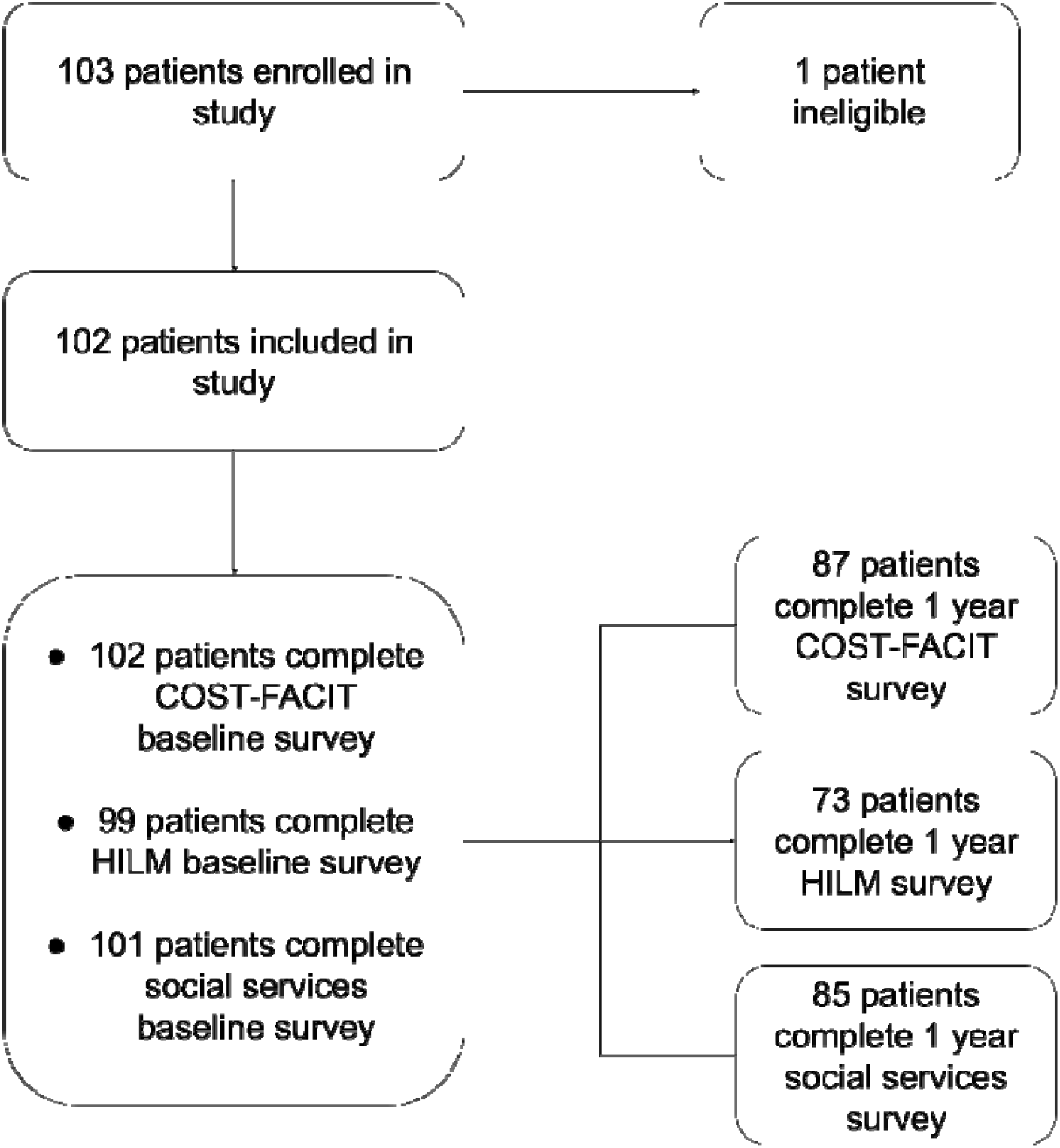
Number of Patients at Beginning and End of Study.

When comparing COST-FACIT scores against time of survey completion, there was an overall statistical difference across the months surveyed (p < 0.001) with omnibus repeated measure F-test. Post hoc contrasts (Tukey-adjusted) found significant differences between baseline versus six months (p = 0.003) and baseline versus one year (p = 0.003). Comparing scores to staging of breast cancer, there was no statistical difference (p = 0.742) across stages. The combined scores with age differed significantly by age (p = 0.001). Scores were significantly higher in the 65+ group at six months (p = 0.018), but not the other time points. Lastly, there was no significant difference (p = 0.817) between insurer and financial toxicity for patients.

## Discussion

From this study, we see that patients had increased COST-FACIT scores throughout the year. This indicates that their level of financial security/confidence increased over time. Interestingly, previous work has shown that the opposite can present in patients with breast cancer. One study found that as patients progressed through treatment, their level of financial burden increased [7]. However, this can be explained with the care approach taken at the Guthrie Cancer Center. Patients seen at the center are offered support including social work, nurse navigation, financial advocacy, and a support group, with assistance available for insurance coverage concerns, transportation issues, and access to various funds that may be able to assist patients with medication costs, household bills and other expenses such as travel. This can alleviate both direct and indirect costs associated with cancer care over time. While the majority did not use and/or were not interested in services provided in the survey, those that utilized it may have found notable improvement in perceived burden. Additionally, one pilot study found that patients express interest in having more information provided early on in the diagnosis and treatment course [12]. By connecting patients early on in treatment, patients at our rural site may have these concerns addressed early on, which could help offset later financial burden that accumulates.

Our work also showed that patients at or over the age of 65 had increased financial stability via COST-FACIT score analysis. This is in line with previous studies conducted that analyze patients of different ages undergoing cancer treatment [4, 13-15]. Explanations for this include the understanding that as people age, they are able to build up more savings that can be used to cushion the financial burden from cancer diagnosis and treatment [14]. Furthermore, older patients may be retired with a set income, or have been employed for a number of years and have greater job security compared to younger adults starting in the workforce [14, 16]. It is possible that Medicare eligibility may play a role in stability. However, this is difficult to ascertain as patients may have different plans within Medicare, thus affecting possible coverage. This work adds to the growing body of literature that more resources need to be provided to the younger patient population as they undergo breast cancer treatment, including in a rural setting.

COST-FACIT survey analysis also demonstrated no significant difference across stages. However, prior work has shown significantly higher costs at later stages [17]. This can be explained via a number of routes. Our study, while including patients with stage 3 and 4 diseases, had over twice as many patients in stages 1 and 2. Including additional patients with later-stage diagnoses may lead to a statistically significant difference. Additionally, patient populations at Guthrie may have greater support than those studied in previous work. Finally, at later stages, patients in our study may have elected to undergo cheaper treatment routes. However, this data is unavailable in our current study.

Finally, no significant difference was noted in insurance types. However, while no statistical difference was found, this does not indicate no burden was found. Both publicly and privately insured patients still faced financial toxicity that should be addressed. Our data indicates however, that further work should focus on more individual characteristics such as age and time from diagnosis, rather than insurer.

This work has provided insight into patients in a rural setting undergoing breast cancer treatment. Its strengths include targeting a rural population and investigating timing, age, stage, and insurer characteristics against financial toxicity. Additionally, the European Society for Medical Oncology – Guidance for Reporting Oncology Real-World Evidence was utilized to guide preparation of this manuscript [18]. There are, however, some limitations to our study. This work was limited to a patient population covered in one hospital system. Future studies should be done to compare differences in financial burden alleviating strategies among different hospital systems. Furthermore, our study had over twice as many patients with early-stage diagnoses compared to later stages. A larger study should be performed to compare later stage diagnosis financial burden in a rural setting, compared to earlier stage. Future work should also investigate breast cancer financial toxicity in rural versus urban settings. Additionally, some patients did not complete different surveys at different time points. Reasons for noncompletion were partially tracked during the survey and included withdrawal from the study, relocation, and passing away. Lack of complete surveys for all time points may provide a less complete understanding of the individual patient experience during treatment. Overall, from our work, we see that financial alleviating strategies for rural populations should target younger patients under 65, as well as providing support and financial strategies early in the diagnosis of patients with breast cancer.

## Conclusion

From evaluating the COST-FACIT score of breast cancer patients at the Guthrie Cancer Center, we observed a significant decrease in perceived financial toxicity over time. Additionally, patients under the age of 65 demonstrated higher perceived financial toxicity when compared to patients at or over 65. This study shows the need to provide additional support to these groups, especially those in a rural setting. Further exploration should be done on effectiveness of different financial alleviating strategies, larger studies comparing later stage diagnoses in the rural setting compared to earlier stage, as well as comparing financial burden in rural to the urban settings.

## Data Availability

To ensure privacy of patient data, raw data is unavailable.

## Statements and Declarations

### Funding

No funding was utilized for this project.

### Competing Interests

The authors have no competing interests.

### Author Contributions

Cynthia Perry-Keaty created the study concept and design. Jennifer Panek collected and organized the data. Emily Thresher, Christopher D. Manko, and Tyler Zlupko performed data analysis. Christopher D. Manko, Khadija Khan, Tyler Zlupko, Nathan Witteveen, and Apurva Inamdar wrote and edited of the manuscript.

### Data Availability

To ensure privacy of patient data, raw data is unavailable.

### Ethical Approval

This study was approved by The Institutional Review Board of The Guthrie Clinic. The reference number is 1808-42. This study is also registered on ClinicalTrials.gov with the following trial ID: NCT03615573.

### Consent to Participate

Written consent from participants was obtained prior to administration of surveys.

### Consent to Publish

The authors affirm that no individual patient data is presented in this manuscript. As such, individual participant consent for publication was not required.

## Acknowledgements

The authors would like to acknowledge Dr. Burt Cagir for his guidance and mentorship of the project. The authors would also like to thank Vicky L. Hickey for her guidance on the institutional review board and trial registration. The authors would also like to acknowledge Jeremy Albright for his assistance with the statistical analysis of the project.

## Notes

### Competing Interest Statement

The authors have declared no competing interest.

### Author Declarations

The Institutional Review Board of The Guthrie Clinic gave ethical approval for this work. The reference number is 1808-42.

